# Reduction in abdominal symptoms (CFAbd-Score), faecal M2-pyruvate-kinase and Calprotectin over one year of treatment with Elexacaftor-Tezacaftor-Ivacaftor in people with CF aged ≥12 years – The RECOVER study

**DOI:** 10.1101/2023.07.10.23292435

**Authors:** Jochen G. Mainz, Karen Lester, Basil Elnazir, Michael Williamson, Ed McKone, Des Cox, Barry Linnane, Carlos Zagoya, Franziska Duckstein, Anton Barucha, Jane C. Davies, Paul McNally, RECOVER Study Group

## Abstract

**Background:** RECOVER is a multicentre post-approval study of Elexacaftor/Tezacaftor/Ivacaftor (ETI) in pwCF in Ireland and the UK. The CFAbd-Score is the first validated CF-specific patient reported outcome measure (PROM) focusing on gastrointestinal symptoms; it comprises 28 items in 5 domains. In a preliminary study, we previously reported reductions in abdominal symptoms (AS) in pwCF after 26 weeks of ETI-therapy using the CFAbd-Score.

Aim: to assess changes in AS in a second, large cohort and explore novel GI-biomarkers of gut inflammation and cell-proliferation in pwCF over one year of ETI-therapy.

**Methods:** Participants were recruited as part of the RECOVER study at 8 sites (Ireland&UK). The CFAbd-Score was administered prior to ETI-initiation, and subsequently at 1,2,6 and 12 months on treatment. Faecal M2-pyruvate kinase (M2-PK) and calprotectin (FC) were quantified in samples collected at baseline, 1 and 6 months.

**Results:** 108 CFAbd-Scores and 73 stool samples were collected at baseline. After 12 months of ETI-therapy, total CFAbd-Scores had significantly declined (15.0±1.4→9.8±1.2pts/p<0.001), and so had all its five domains of “pain” (16.9±2.0pts→9.9±1.8pts/p<0.01), “GERD” (14.4±1.8→9.9±1.6/p<0.05), “disorders of bowel movements” (19.2±1.4→14.1±1.5/p<0.01), “appetite” (7.0±1.1→4.6±1.2/p<0.01) and “impaired-QoL” (13.3±1.9→7.5±1.5/p<0.001). Levels of M2-PK and FC significantly decreased during ETI-therapy.

**Discussion:** In-depth analysis of AS with the CFAbd-Score reveals a statistically significant, clinically relevant and sustained improvement with ETI. We attribute this to high sensitivity of the implemented CF-specific PROM, developed and validated following FDA-guidelines.

Furthermore, for the first time during ETI-therapy a significant decline in faecal M2-PK, a marker of inflammation and cell-proliferation, was found, in parallel to FC.

**Graphical Abstract:** 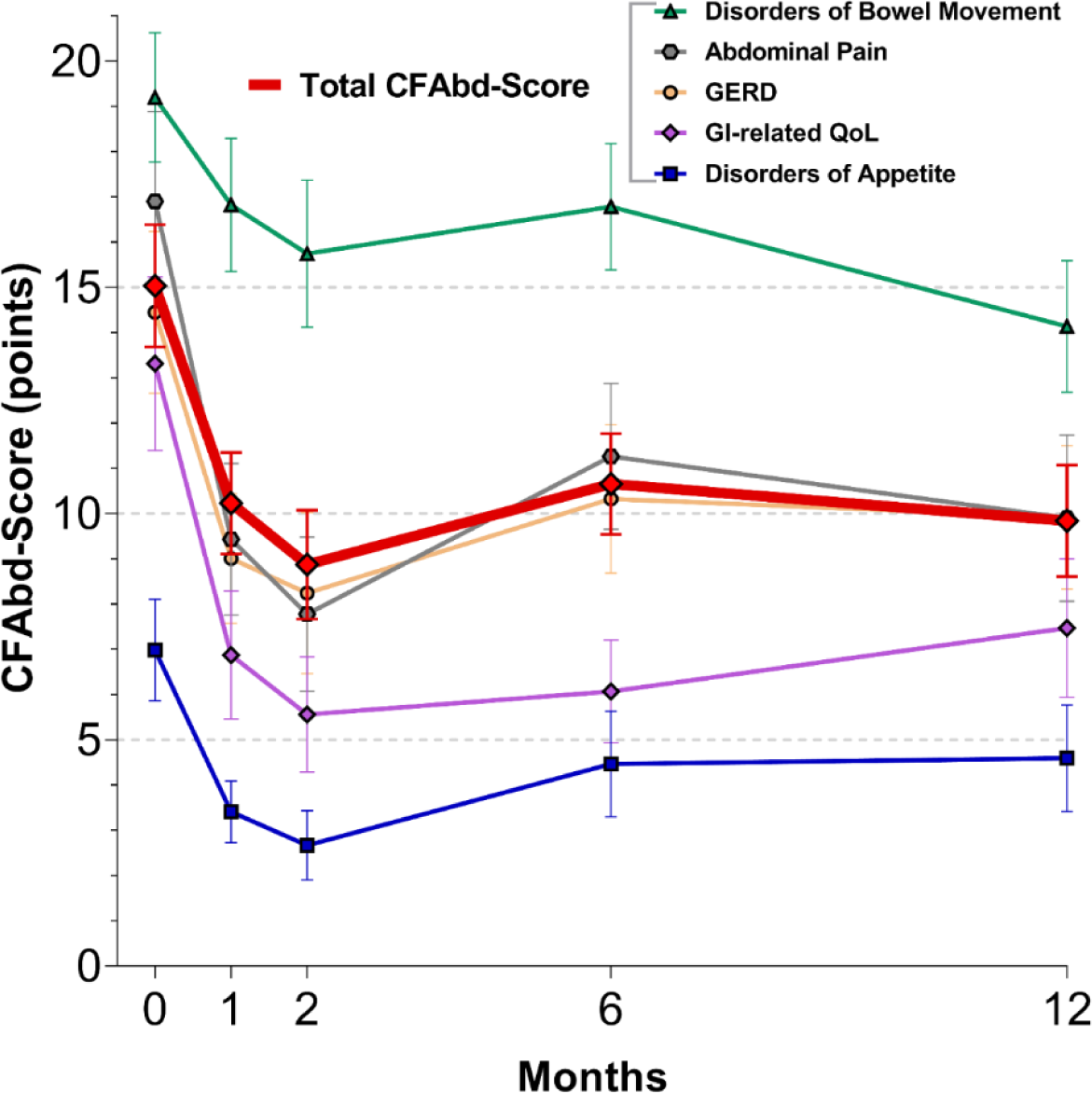

Statistically significant and clinically relevant sustained decline in GI symptoms using the CFAbd-Score in pwCF over 12 months of therapy with Elexacaftor-Tezacaftor-Ivacaftor (ETI) as part of the RECOVER study. Changes over time in total CFAbd-Scores as well as of the 5 included domains are shown before (0), as well as 1,2,6 and 12 months after initiation of ETI

**Highlights:**

- The present study uses the first validated CF-specific patient reported outcome measure focusing on gastrointestinal symptoms (CFAbd-Score) to demonstrate that ETI leads to substantial and sustained reduction in GI-symptom burden
- Symptom improvement is rapid, being evident at 1 month, peaking 2 months and stabilising thereafter
- Previous studies assessing effects of ETI on GI-symptoms had generated conflicting results
- For the first time, significant improvements in faecal M2-pyruvate Kinase (M2-PK), a marker of cell proliferation were seen on ETI, in addition to reductions in faecal calprotectin as previously reported

## 1 Introduction

Already at birth, most infants with cystic fibrosis (CF) have exocrine pancreatic insufficiency (PI). Malabsorption, malnutrition and growth failure follow PI, particularly with advancing age and evolving CF lung disease. Intestinal epithelial dysfunction, particularly in the setting of PI, can be associated with inflammation, dysbiosis and dysmotility, partial or complete bowel obstruction either in infancy with meconium ileus (MI) or later in life with constipation or distal intestinal obstruction syndrome (DIOS).(1) The nature of abdominal involvement in people with CF (pwCF) may result in a significant burden of abdominal symptoms (AS) leading to impairment in quality of life.(1-3) AS including reduced appetite, pain, flatulence and abdominal distention are almost universal, and are associated with abnormal imaging findings.(4-7) Faecal calprotectin (FC), an extensively used biomarker for intestinal inflammation, has been found to be markedly elevated in pwCF, compared with healty controls (HC).(8) We have previously reported on elevated levels of FC, interleukin (IL)-1β, M2-pyruvate kinase (M2-PK) and neutrophil elastase (NE) in feces of pwCF compared to HC.(9) Specifically, pwCF reporting a high burden of abdominal pain displayed significantly elevated FC levels. PwCF and their care teams have ranked gastrointestinal presentations of CF as a high research priority in CF.(7,10) Abdominal manifestations, although they can be very troublesome, can be hard to characterize, challenging to effectively treat and receive less attention overall than pulmonary manifestations in pwCF.

The CFAbd-Score, the first CF-specific AS questionnaire and its scoring algorithm which weights different items and domains differently, was designed and validated according to FDA and COSMIN guidelines.(4-6,9,11-13) It was developed in close collaboration with the CF community voice (pwCF, their proxies, as well as CF care-specialist of different professions: pediatricians, pulmonologists, gastroenterologists, nutritionalists, psychologists). The initial 5-sided version (JenAbd-Score (22)) was condensed to a one-sided PROM, the CFAbd-Score, including 28 items in 5 domains(4-6) with a 2-week recall period. The score ranges from a minimum of 0 to a maximum of 100 points with higher scores representing a more pronounced symptom burden. CFAbd-Scores correlate with CFTR-genotype and phenotype, as well as with history of abdominal surgery and both intestinal inflammation(9) and imaging findings(5) in pwCF.

In the last decade, the use of CFTR-modulating therapy has grown significantly among pwCF. The first CFTR modulator, ivacaftor (IVA) led to marked improvements in sweat chloride and end-organ function in those with responsive CFTR mutations, representing only a small minority (5%) of pwCF.(14-19) Recently, the triple combination modulator Elexacaftor-Tezacaftor-Ivacaftor (ETI) has demonstrated similar dramatic improvements in sweat chloride and end organ function, but is suitable for use in >90% of pwCF.(20-25) Collectively IVA and ETI are referred to as highly effective CFTR-modulating therapies (HEMT).

In early studies involving HEMT, improvement in the digestive system had only been indirectly captured by the substantial incremental improvement in the nutritional status of pwCF on treatment. As clinical trials of ivacaftor moved into younger ages, improvements in faecal elastase (FE-1) levels (marker of exocrine pancreatic insufficiency) in some cases were seen in the normal range.(14,17) This has been reproduced in real-world studies.(26,27) Use of ivacaftor has also been associated with improvements in gastric hyperacidity, markers of intestinal inflammation,(28) and the gut microbiome.(29) In a similar vein, ETI has been associated with improvements in FE-1 in younger children(30) and in gastrointestinal inflammation in older children and adults.(31)

While HEMT has been repeatedly shown to improve overall quality of life in CF as measured by CFQ-R,(20,32) data on AS have been more sparse, using different measures and producing differing results.(13,31) Using previously developed (non-CF) AS questionnaires like the PAGI-SYM, PAC-SYM, PAC-QoL or Peds-QL, as well as questionnaires developed for irritable bowel syndrome(31,33,34) and the abdominal domain of CFQ-R,(35) a high burden of GI symptoms in pwCF has been demonstrated. However, in a recent large post-approval study of ETI in the US including 267 pwCF, although changes in these symptom scores (PAGI-SYM, PAC-SYM, PAC-QoL) were statistically significant, they were not of a magnitude considered clinically relevant.(31) Also recently, a smaller single center retrospective observational study implemented the CFQ-R PROM to assess changes in GI symptoms in pwCF during ETI. However, no improvement in the subdomains regarding GI symptoms of “Eating Problems”, “Weight” and “Digestive Problems” was observed.(36)”

In contrast, using the CF-specific CFAbd-Score, we have previously demonstrated statistically and clinically significant improvements in AS in 107 pwCF aged ≥12 years from Germany (n=68) and the UK (n=39) over 26 weeks.(13) This study was limited in some aspects owing to marked differences in demographic features between German and UK populations with regard to age, *CFTR* mutations, exocrine pancreatic status, and recruitment time frame. While these issues were recognised by the authors, the need for a replicate study over a longer time period, linked to other gastro-intestinal outcomes was evident, hence the design and conduct of this aspect of the RECOVER study.

RECOVER (NCT04602468) is a multi-centre post approval study designed to examine the impact of ETI on a range of clinically relevant outcome measures in a real-world setting in pwCF in Ireland and the UK.

Our aim in this study was to characterise the extent and timing of changes to abdominal symptoms and markers of intestinal inflammation and cell proliferation among pwCF starting treatment with ETI.

## 2 Methods

### 2.1 Participants

As part of the RECOVER study, participants who were either homozygous for the F508del mutation (FF) or heterozygous for F508del and a minimum function mutation (FMF) were enrolled at 8 clinical sites across Ireland and the UK prior to initiation of ETI as part of routine care. Data on baseline characteristics, and longitudinal data on height, weight and BMI, collected every 3 months, were taken from medical notes. Research Ethics Committees approved the study and informed consent and assent was obtained.

### 2.2 Symptoms assessment - CFAbd-Score

The CFAbd-Score, a PROM consisting of 28 questions in 5 domains, was applied prior to ETI-initiation, and subsequently at 1,2,6 and 12 months on treatment. Total CFAbd-Scores and domain scores range from 0 to 100 points, with higher scores indicating higher AS-burden.(6) Total CFAbd-Scores and domain scores are calculated using a scoring algorithm that weights items and domains differently to optimize sensitivity, as published elsewhere.(6,13) Detailed information about items and domains of the CFAbd-Score is attached as supplement. CFAbd-Score questionnaires were administered to participants by research staff at sites. Pseudonymized questionnaires were captured, processed and scored centrally at the CF Center in Brandenburg and der Havel, Germany.

### 2.3 Collection and processing of stool specimens

Stool specimens were collected at baseline and one and six months after initiation of ETI therapy. Participants were provided in advance with equipment to collect and refrigerate stool samples on the morning of each study visit. After collection, stool samples were split at the study site. 5-10g aliquots were frozen at −80C, with one aliquot sent on dry ice to the CF Center in Brandenburg an der Havel, Germany for analysis of calprotectin, M2-PK and FE-1. Further aliquots were frozen at −80C and shipped to central labs at Children’s Health Ireland and stored for future microbiome and other analysis.

### 2.4 Markers of faecal inflammation

Calprotectin was measured from frozen stool samples using the DiaSorin LIAISON® Calprotectin assay (DiaSorin Inc. Stillwater, MN 55082, US), a sandwich-test using two monoclonal antibodies, an in-vitro-diagnostic chemiluminescent immunoassay (CLIA), range 5-800µg/g; Liaison®XL. M2-PK was measured using the ScheBo Tumor M2-PK kit (ScheBo Biotech, Giessen, Germany), Reader ELx800TM (Fa.BioTek) an ELISA with a range from 1-20U/ml. Stools were extracted into ready-to-use extraction buffer (anti-bio-POD-step complex) using the ScheBo® Master Quick-Prep™ and each 50µl of stool extract was analyzed. FE-1 was measured with a sandwich-test using two monoclonal antibodies, an in-vitro-diagnostic chemiluminescent immunoassay (CLIA)(DiaSorin LIAISON® Elastase-1 Assay (DiaSorin Inc. Stillwater, MN 55082Calpro, San Diego, CA, US). Range: 0.2µg/g-800µg/g stool, Liaison^®^XL.

### 2.5 Statistical analysis

Linear mixed-effects models for repeated measures were used to test for differences in the CFAbd-Score and its five domains, including time from therapy start (in months) as fixed effect as well as variables sex (M/F), previous CFTR-modulating therapy (yes/no), genotype (F508del-homozygous/heterozygous) and group age (<18/≥18) as covariates. Significant first-order interactions of these covariates and time were selected using backward-elimination procedures. Total CFAbd-Score and domains were log-transformed to satisfy normality conditions on residuals. Results are reported as means±error of mean (SEM) in the original scale. For between-group (cross-sectional) comparisons of FC, FE-1 and M2-PK all data available at each time-point was used. For FE-1 and M2-PK longitudinal comparisons, the non-parametric Wilcoxon signed-rank test for paired samples was used; FE-1 concentrations from pwCF identified as pancreatic sufficient (PS) at baseline were excluded from the longitudinal FE-1 comparisons. As normal reference values for FC, M2-PK and FE-1 concentrations, we considered the cutoff values of 50µg/g, 4U/ml and 200µg/g, respectively.(8,37,38)

## 3 Results

### 3.1 Participants

A total of 117 pwCF were recruited to the 12 and over age cohort of the RECOVER study, 108 of whom completed the CFAbd-Score at baseline and at least one other time point. Demographics and clinical information is shown in Table 1.

**Table 1.**
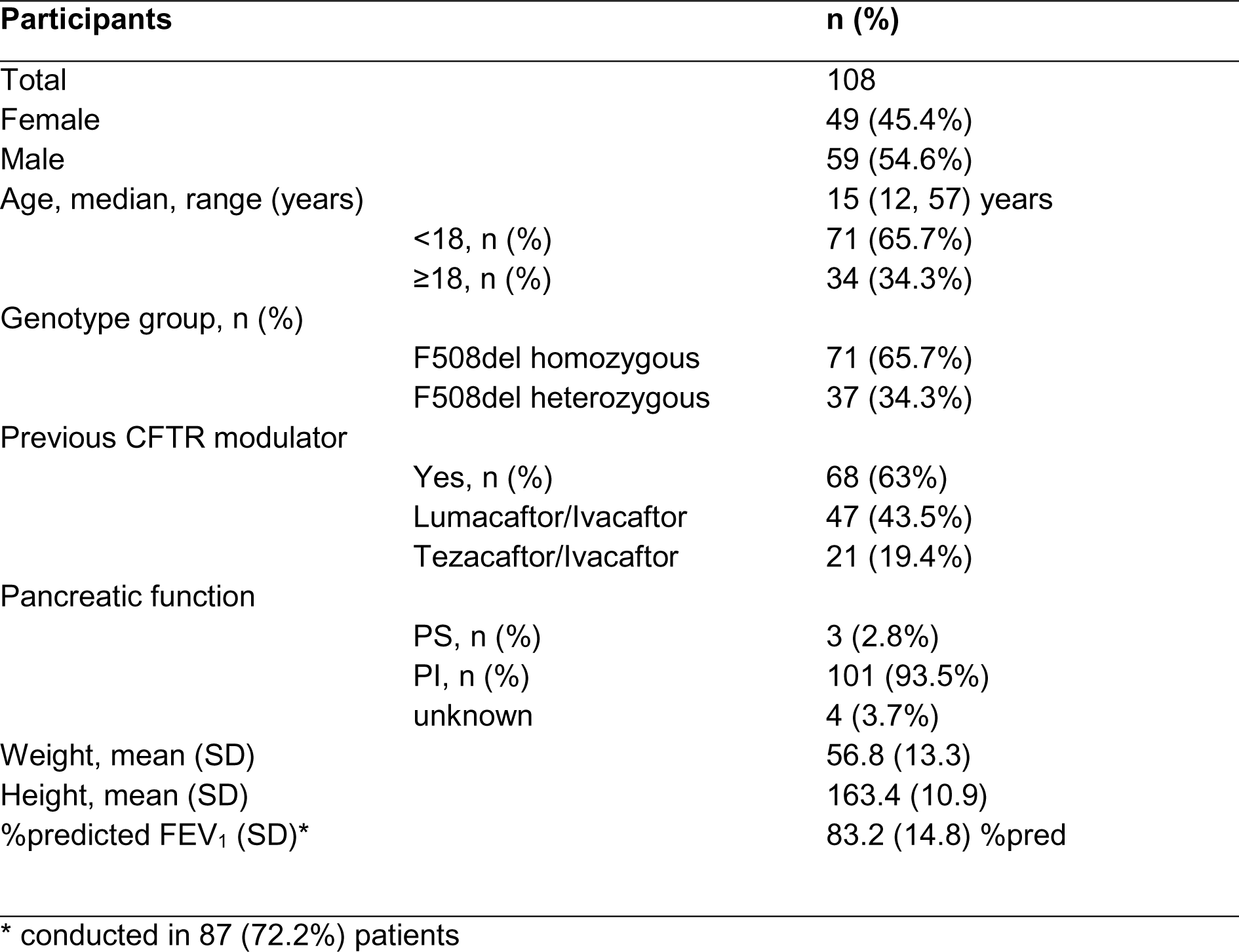
Demographic and clinical information of participants included in the study.

### 3.2 CFAbd-Score

Total CFAbd-Scores improved significantly from baseline over one year of treatment, as did subscores for “pain”, “GERD”, “disorders of appetite” (DA), “QoL” and “disorders of bowel movements” (Fig.1-Table 2). Improvements are rapid, with the major improvements occurring in the first month and attaining a maximum improvement at 2 months for total scores and most of the domains (Fig.2).

**Figure 1.**
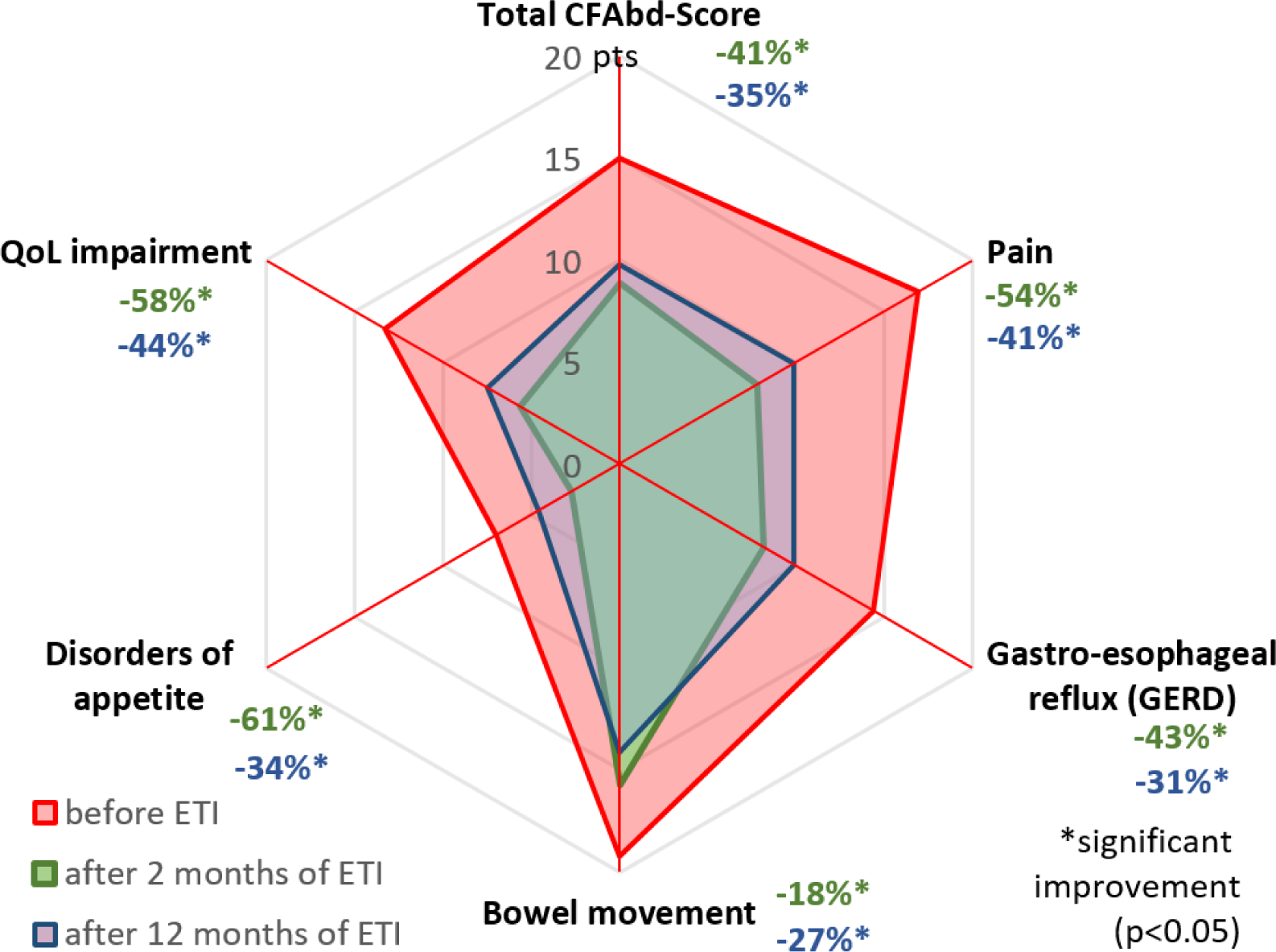
Baseline (solid red line), 2-month (green line) and 12-month (blue line) means of total CFAbd-Score and its five domains, i.e. pain, gastroesophageal reflux disease (GERD), disorders of bowel movement (DBM), disorders of appetite (DA) and impairment of quality of life (QoL).

**Figure 2.**
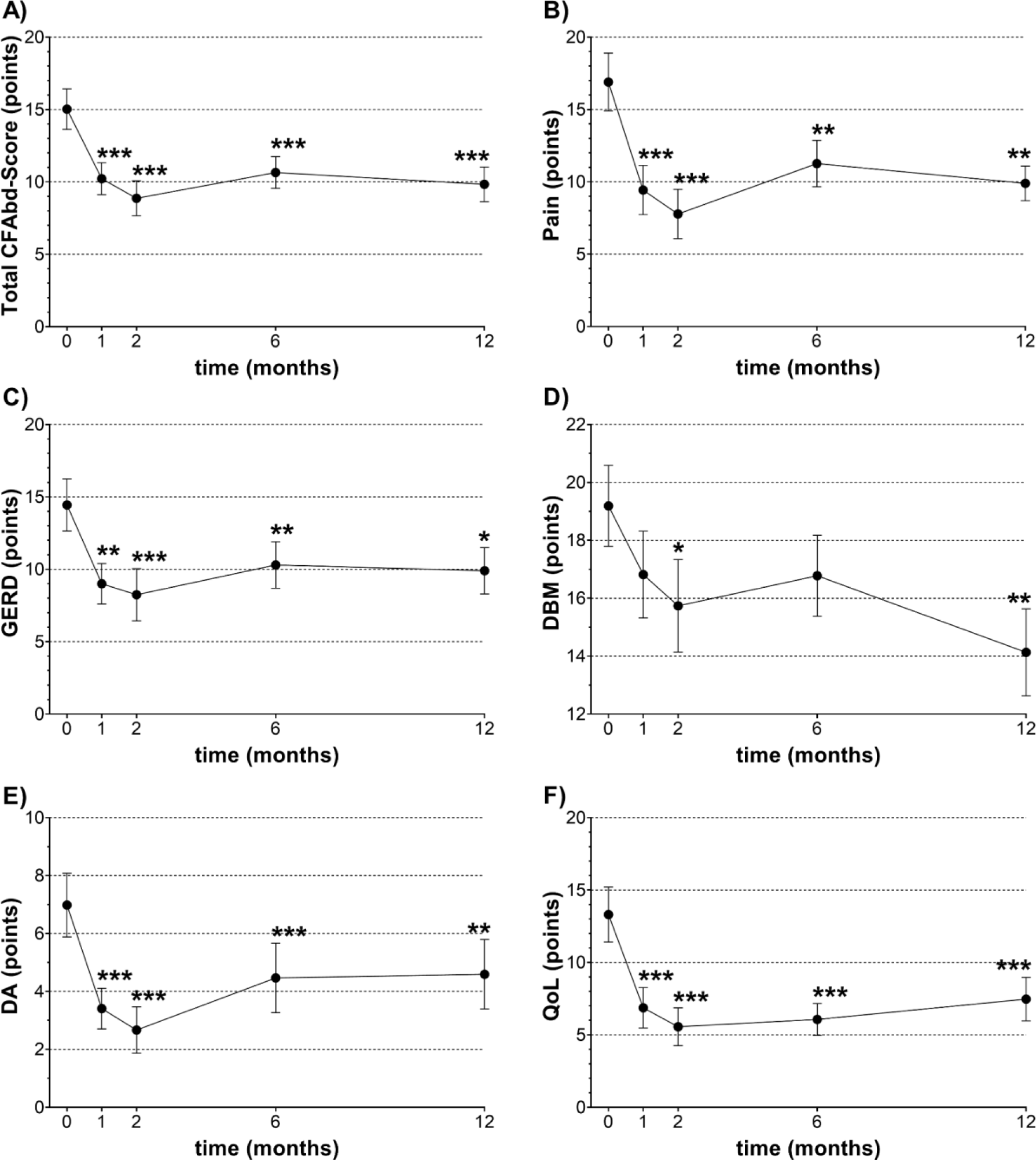
Mean scores (mean±SEM) at baseline (n=108) and after one (n=97), two (n=72), six (n=103) and 12 (n=81) months of ETI therapy for: A) total CFAbd-Score, B) pain domain, C) gastroesophageal reflux disease (GERD) domain, D) disorders of bowel movement (DBM) domain, E) disorders of appetite (DA) domain and F) impairment of quality of life (QoL) domain. Significant changes with respect to baseline values are labelled as * p<0.05, ** p<0.01, *** p<0.001.

**Table 2.** Changes in means (mean±SEM) of total CFAbd-Score and its five domains, i.e. pain, gastroesophageal reflux disease (GERD), disorders of bowel movement (DBM), disorders of appetite (DA) and impairment of quality of life (QoL).

|  | Month |  |  |  |  |
| --- | --- | --- | --- | --- | --- |
|  | 0<br>n =108 | 1<br>n=97 | 2<br>n=72 | 6<br>n=103 | 12<br>n=81 |
| **Total CFAbd-Score** | 15.0±1.4 | 10.2±1.1*** | 8.9±1.2*** | 10.7±1.1*** | 9.8±1.2*** |
| **Pain** | 16.9±2.0 | 9.4±1.7*** | 7.8±1.7*** | 11.3±1.6** | 9.9±1.8** |
| **GERD** | 14.4±1.8 | 9.0±1.4** | 8.2±1.8*** | 10.3±1.6** | 9.9±1.6* |
| **DBM** | 19.2±1.4 | 16.8±1.5 | 15.7±1.6* | 16.8±1.4 | 14.1±1.5** |
| **DA** | 7.0±1.1 | 3.4±0.7*** | 2.7±0.8*** | 4.5±1.2*** | 4.6±1.2** |
| **QoL** | 13.3±1.9 | 6.9±1.4*** | 5.6±1.3*** | 6.1±1.1*** | 7.5±1.5*** |

Significant differences with respect to baseline (month 0) values are labelled as \* p<0.05, \*\* p<0.01, \*\*\* p<0.001.

Overall, in the “GERD” domain, adult pwCF (age≥18) reported significantly higher scores than adolescents (baseline [mean±SEM]: 16.2±2.9 vs 13.1±2.3, 12-months: 15.2±3.2 vs 7.0±1.8; main-effect-p=0.003). Females reported significantly higher scores than males in the “DA” domain (baseline [mean±SEM]: 4.5±0.1 vs 10.0±0.3, 12-months: 7.9±0.4 vs 2.2±0.1; main-effect-p=0.01). No other differences regarding sex, previous CFTR-modulator therapy, genotype group or age (<18/≥18) were observed.

### 3.3 Faecal markers of inflammation and cell proliferation

Overall, 93 participants provided at least one stool sample for analysis of FC, FE-1 and M2-PK. Specifically, stool samples from 73, 74 and 62 patients were collected at baseline, 1 month and 6 months, respectively. For analysis of FC changes over the six-month observation time frame, a linear mixed-effects model was used including all available samples at each time point. For M2-PK and FE-1, however, only data from pwCF that provided stool samples at all three time points was analysed (n=43 for M2-PK and n=42 for FE-1).

FC concentrations decreased significantly from baseline to 1 month (mean±SEM) (70±8.9µg/g→27.4±3.4µg/g/p<0.0001). This difference was maintained at 6 months (27.3±3.4µg/g). In parallel, median M2-PK-concentration decreased significantly by 76% from baseline to 1 month (median [IQR]): 12.0U/ml [4.4-20.0]→2.9U/ml [1.0-8.0]/p<0.0001), with little change thereafter (median [IQR]: 2.7U/ml [1.0-10.7])(Fig.3). There was a strong correlation between FC and M2-PK at baseline (Spearman’s rho=0.50/p<0.0001, n=73). This correlation was weaker at 6 months (Pearson’s r=0.34/p<0.05, n=61).

**Figure 3.**
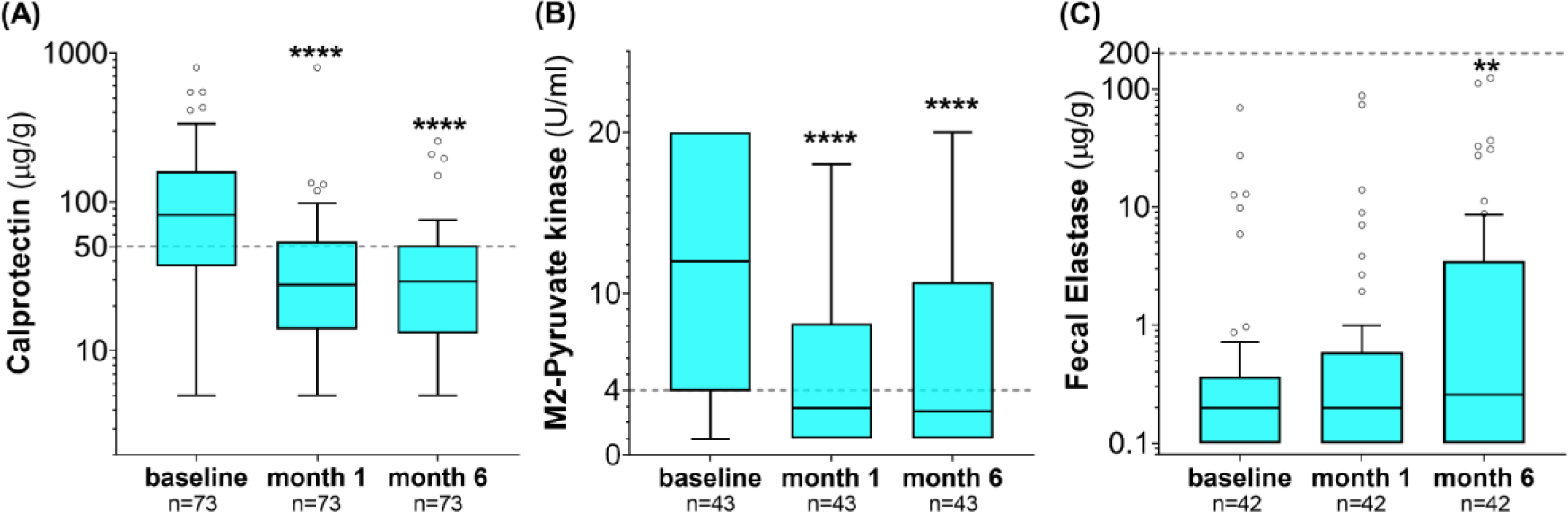
Values at baseline, month 1 and month six for (A) faecal calprotectin (n=73 (baseline), 74 (month 1) and 62 (month 6)), (B) M2-pyruvate kinase (n=43) and (C) faecal elastase (n=42). Horizontal lines in boxes of all boxplots depict median values. FC mean values and SEM resulting from the linear mixed-effects model are: 70±8.9 (baseline), 27.4±3.4 (1 month) and 27.3±3.4 (6 months). Dashed lines represent the corresponding normal reference values for each parameter. P values are labelled as ** p<0.01 and **** p<0.0001 compared to baseline values.

### 3.4 Markers of pancreatic sufficiency

At baseline, 87.5% (63/72) of participants had levels of FE-1 activity <5µg/g, indicative of severe pancreatic dysfunction. At 6 months, 85% (51/60) of participants had levels <5µg/g. While median FE-1 concentrations were significantly higher at 6 months compared to baseline (median [IQR]: 0.3µg/g[0.1-3.1], 0.2µg/g[0.1-0.4]/p=0.003), this was not at a level where clinical effects would be apparent. 2/60 participants had FE-1 concentrations >100µg/g (123µg/g and 111µg/g) at 6 months.

### 3.5 Inflammatory and proliferation-parameters and correlations with abdominal symptoms

At baseline, a significant positive correlation between FC and both the “Disorders of Appetite” (Pearson’s r=0.32/p=0.005) and the “QoL” (Pearson’s r=0.29/p=0.01) domains were observed. Baseline M2-PK concentrations correlated positively with baseline total CFAbd-Score (Spearman’s rho=0.31/p=0.001) and the “Pain” domain (Pearson’s r=0.32/p=0.01). FE-1 did not correlate with the CFAbd-Score or any of its five domains.

At month six, there were no correlations with FC. M2-PK concentrations still correlated with total CFAbd-Scores (Spearman’s rho=0.31/p=0.02), as well as now with “GERD” scores (Spearman’s rho=0.31/p=0.02). Furthermore, “QoL” domain scores positively correlated with FE-1 concentration (Pearson’s r=0.29/p=0.03).

## 4 Discussion

Previously, intestinal manifestations did not receive much attention due to the intrinsic complexity in their characterization and treatment. Nowadays, intestinal manifestations in pwCF are acknowledged to result in a high burden of symptoms and their resolution was listed as second most important of the 10 top research priorities in CF.(4,7,9,10) Nevertheless, although the use of highly effective CFTR-modulating therapy including ivacaftor and ETI has been shown to improve general Quality of life and GI manifestations in pwCF,(17,28,39) the use of different PROMs to assess changes in AS led to conflicting results.(13,31) Furthermore, data on ETI effects on gastrointestinal manifestations and their link to other outcomes, e.g. markers in stool, is still scant.

The aim of this study was to characterise the extent and timing of changes to AS, intestinal inflammation and cell proliferation markers among pwCF starting ETI-treatment. Accordingly, the present study fills the need for a thorough GI-symptom assessment in a prospective study including a homogeneous and very well controlled cohort of pwCF receiving ETI, including series of consecutive time points, linked with markers of intestinal inflammation and cell proliferation.

Early results obtained with the CFAbd-Score in a longitudinal study, including 107 pwCF aged ≥12 years from Germany and the UK receiving ETI treatment over a period of 26 weeks,(13) had revealed marked declines for the total PROM (−29%) and its five domains. However, although the reduction in symptoms ranging between −23% and −67% was clearly within the margin quoted as clinically significant, changes in CFAbd-Scores revealed marked differences between both German and British cohorts. Specifically, improvement in the 68 German pwCF was markedly higher than the improvement in the 39 British pwCF, which was attributed to the marked differences in demographic features of German and UK pwCF, e.g. in regard to age, *CFTR* mutations, PS/PI, and recruiting time frame.

In contrast, the GALAXY study used previously developed (non-CF) AS-questionnaires including the PAGI-SYM, PAC-SYM, PAC-QoL, as well as a modified Bristol stool scale.(31,33,34) Although the reported changes in GI symptoms in 267 pwCF followed up before and during ETI therapy were statistically significant, according to the authors, they lay below the margin of clinical significance.(31) Furthermore, recently a smaller single center retrospective observational study assessed changes in CFQ-R scores during ETI.(36) In contrast to other CFQ-R subdomains, no improvement in the GI symptoms of “Eating Problems”, “Weight” and “Digestive Problems” was observed.

As part of the RECOVER study, we have thoroughly investigated longitudinal changes in AS assessed with the CF-specific CFAbd-Score from 108 pwCF aged 12 years and older. The total CFAbd-Score, measured at baseline, 1, 2, 6, and 12 months after ETI initiation declined by 41% after 2 months, which for the total CFAbd-Score and four domains accorded to the lowest AS-burden. Thereafter, and up to 12 months of observation, a slightly increasing trend was seen in some domains, which remained significantly lower than baseline at 12 months. Except for “Disorders of Bowel Movement”, scores for all 5 domains were significantly lower than baseline at each observed time point.

Similarly to previous publications regarding the impact of HEMT on inflamation in pwCF,(31,40) FC in this study decreased after 1, 2 and 6 months of therapy and a significant correlation between baseline FC and the “Pain” domain in the CFAbd-Score was found. This result concurs with a previous cross-sectional study with the CFAbd-Score, wherein FC was found to be elevated in pwCF suffering from a high burden of pain.(9) This finding additionally corresponds with studies revealing a HEMT-induced decrease in inflammation in pwCF in different organ systems such as the lower- and upper-airway.(41,42)

Unlike the study by Schwarzenberg et al.,(31) during ETI therapy, FE-1 concentrations appeared to significantly increase in our cohort. However, this increase occurred in only a subset of participants and did not reach a level likely to be of clinical relevance. After 6 months of ETI, only 2/60 pwCF had FE-1 values above 100µg/g, but below the pancreatic sufficiency range considered to be above 200µg/g. This contrasts with the response observed for ivacaftor in young age cohorts, by which pwCF with a gating mutation maintain or even re-gain some degree of pancreatic sufficiency.(38,43,44)

To our knowledge, this is the first time that a decrease in faecal M2-PK associated with the use of ETI has been reported. The dimeric PK, elevated in 32/43 (74.4%) of the study participants with a median at baseline of 12U/ml (normal: <4.0U/ml(37)), declined significantly after one month, and remained low after six months of therapy (2.9 and 2.7U/ml, respectively). This finding may be of particular relevance, as M2-PK has not been deemed to directly represent inflammation.(45-48) Instead, it reflects epithelial turnover and it has been reported to be expressed in most human tumours. Herein, it is deemed to play a central role in metabolic reprogramming of cancer cells being essential in their indiscriminate proliferation, survival, and tackling apoptosis.(49) Correspondingly, an increased epithelial turnover has also been linked to elevated rates of GI malignancies in pwCF and, consequently, elevated levels of M2-PK in stool of pwCF may be hypothesized to be correlated with their higher risks of colorectal cancer.(50) Interestingly, a number of studies are presently examining anti-M2-PK therapies aiming at reducing cancer risks by inhibiting the mediators’ putative cell-proliferating properties.(49,51)

Several authors have recently projected that the improved survival in pwCF together with the higher predisposition and earlier onset of CFTR-associated intestinal malignancies will lead to higher cancer incidences in this cohort.(28,43,52) In contrast, this novel finding of reduced faecal M2-PK with ETI treatment could be a notable finding, perhaps suggesting down-regulation of carcinogenic effects of CFTR-deficiency. Such fundamental roles of CFTR in cellular processes such as foetal development, epithelial differentiation/polarization, and regeneration, as well as in epithelial–mesenchymal transition (EMT) have recently been reviewed by Amaral et al.,(53) wherein the authors address dysfunctional CFTR as a possible promotor of cancer progression. Re-constitution of CFTR function by ETI could therefore be hypothesized to have effects of a tumor suppressor. Further work will be needed to establish this.

Limitations of this study may include ceiling and floor effects arising in M2-PK laboratory analyses associated with measurement ranges of the assays used in this study (Fig. 3). However, although tests involving wider measurement ranges are commonly preferred, wider ranges in this study are unlikely to have a large effect on the conclusions reached herein.

This can be seen for instance in Fig.3B, where about 30% (13/43) of all M2-PK samples at baseline reached the maximum of the measurement scale, whereas only 9% (4/43) of the samples reached the minimum of the scale. Conversely, at six months about 35% (15/43) of all M2-PK samples reached the minimum level achieved by the assay, and only 9% (4/43) resulted above the maximum range value. Consequently, an M2-PK test covering wider measurement ranges would have little to no effect on median values at each time point and, similarly, we would expect little effect on the corresponding p-values. However, a possible effect would be an even more pronounced M2-PK normalization in pwCF treated with ETI than in the present results.

## Conclusion

This study adds to the growing evidence that ETI reduces GI-symptoms in pwCF, highly significant findings being likely due to the sensitivity of the CF-specific CFAbd-Score used to measure symptoms in this study. GI symptoms seem to be at their minimum at two months after treatment initiation. Similar to others, we have confirmed a significant improvement in gut inflammation with ETI use in pwCF, as measured by FC.

Finally, to the best of our knowledge for the first time, we demonstrate a significant improvement with ETI in faecal levels of M2-PK, a marker of cell turnover and a biomarker for colorectal carcinoma.

Collection of further longitudinal data and biological samples as part of the RECOVER study is ongoing.

## Data Availability

All data produced in the present study are available upon reasonable request to the authors

## Acknowledgements

We would like to acknowledge the dedication and commitment of people with CF and their parents who participated in this study, the CF care teams at the study sites, the RECOVER site staff and coordinating team. Funding for this study was from Cystic Fibrosis Foundation (MCNALL19K0), CF Trust (VIA118) and CF Ireland.

JCD is supported by the National Institutes of Health & Care Research through the Imperial Biomedical Research Centre, the Brompton Clinical Trials Facility and a Senior Investigator Award. The site is supported as part of the CF Trust’s Clinical Trials Accelerator Platfor, (CTAP).

Furthermore, we would like to acknowledge Karina Börner and Yvonne Schimpf (Brandenburg an der Havel) for their assistance in laboratory.

## CRediT authorship contribution statement

JGM: Conceptualization, Methodology, Stool analysis, Providing and analysing CFAbd-Scores, Writing – Visualization, original draft, review & editing

KL Project administration, Formal analysis, Data curation

BE Sample and data collection, local supervision

MW Sample and data collection, local supervision

EMK Sample and data collection, local supervision

DC Sample and data collection, local supervision

BL Sample and data collection, local supervision

CZ Stool Analyses, Providing and analysing

CFAbd-Scores, Data curation, Data analysis, Visualization, Writing – original draft, review & editing

FD Writing – review & editing

AB Stool Analyses, Writing – review & editing

JCD Conceptualization, Methodology, Formal analysis, Writing – review & editing, Supervision, Funding acquisition

PMN Conceptualization, Methodology, Formal analysis, Project administration, Supervision, Writing – review & editing, Funding acquisition

## References

1. Sathe MN, Freeman AJ. Gastrointestinal, Pancreatic, and Hepatobiliary Manifestations of Cystic Fibrosis. Pediatr Clin North Am. 2016;63(4):679–98.

2. de Freitas MB, Moreira EAM, Tomio C, Moreno YMF, Daltoe FP, Barbosa E, et al. Altered intestinal microbiota composition, antibiotic therapy and intestinal inflammation in children and adolescents with cystic fibrosis. PLoS One. 2018;13(6):e0198457.

3. Dorsey J, Gonska T. Bacterial overgrowth, dysbiosis, inflammation, and dysmotility in the Cystic Fibrosis intestine. J Cyst Fibros. 2017;16 Suppl 2:S14–S23.

4. Tabori H, Arnold C, Jaudszus A, Mentzel HJ, Renz DM, Reinsch S, et al. Abdominal symptoms in cystic fibrosis and their relation to genotype, history, clinical and laboratory findings. PLoS One. 2017;12(5):e0174463.

5. Tabori H, Jaudszus A, Arnold C, Mentzel HJ, Lorenz M, Michl RK, et al. Relation of Ultrasound Findings and Abdominal Symptoms obtained with the CFAbd-Score in Cystic Fibrosis Patients. Sci Rep. 2017;7(1):17465.

6. Jaudszus A, Zeman E, Jans T, Pfeifer E, Tabori H, Arnold C, et al. Validity and Reliability of a Novel Multimodal Questionnaire for the Assessment of Abdominal Symptoms in People with Cystic Fibrosis (CFAbd-Score). Patient. 2019;12(4):419–28.

7. Moshiree B, Freeman AJ, Vu PT, Khan U, Ufret-Vincenty C, Heltshe SL, et al. Multicenter prospective study showing a high gastrointestinal symptom burden in cystic fibrosis. J Cyst Fibros. 2022.

8. Ellemunter H, Engelhardt A, Schuller K, Steinkamp G. Fecal Calprotectin in Cystic Fibrosis and Its Relation to Disease Parameters: A Longitudinal Analysis for 12 Years. J Pediatr Gastroenterol Nutr. 2017;65(4):438–42.

9. Jaudszus A, Pfeifer E, Lorenz M, Beiersdorf N, Hipler UC, Zagoya C, et al. Abdominal Symptoms Assessed With the CFAbd-Score are Associated With Intestinal Inflammation in Patients With Cystic Fibrosis. J Pediatr Gastroenterol Nutr. 2022;74(3):355–60.

10. Rowbotham NJ, Smith S, Leighton PA, Rayner OC, Gathercole K, Elliott ZC, et al. The top 10 research priorities in cystic fibrosis developed by a partnership between people with CF and healthcare providers. Thorax. 2018;73(4):388–90.

11. Administration USDoHaHSFaD. GUIDANCE DOCUMENT: Patient-Reported Outcome Measures: Use in Medical Product Development to Support Labeling Claims 2009 10/17/2019.

12. Mokkink LB, Terwee CB, Patrick DL, Alonso J, Stratford PW, Knol DL, et al. The COSMIN study reached international consensus on taxonomy, terminology, and definitions of measurement properties for health-related patient-reported outcomes. J Clin Epidemiol. 2010;63(7):737–45.

13. Mainz JG, Zagoya C, Polte L, Naehrlich L, Sasse L, Eickmeier O, et al. Elexacaftor-Tezacaftor-Ivacaftor Treatment Reduces Abdominal Symptoms in Cystic Fibrosis-Early results Obtained With the CF-Specific CFAbd-Score. Front Pharmacol. 2022;13:877118.

14. Davies JC, Cunningham S, Harris WT, Lapey A, Regelmann WE, Sawicki GS, et al. Safety, pharmacokinetics, and pharmacodynamics of ivacaftor in patients aged 2-5 years with cystic fibrosis and a CFTR gating mutation (KIWI): an open-label, single-arm study. Lancet Respir Med. 2016;4(2):107–15.

15. Davies JC, Wainwright CE, Canny GJ, Chilvers MA, Howenstine MS, Munck A, et al. Efficacy and safety of ivacaftor in patients aged 6 to 11 years with cystic fibrosis with a G551D mutation. Am J Respir Crit Care Med. 2013;187(11):1219–25.

16. Davies JC, Wainwright CE, Canny GJ, Chilvers MA, Howenstine MS, Munck A, et al. Efficacy and safety of ivacaftor in patients aged 6 to 11 years with cystic fibrosis with a G551D mutation. American journal of respiratory and critical care medicine. 2013;187(11):1219–25.

17. Davies JC, Wainwright CE, Sawicki GS, Higgins MN, Campbell D, Harris C, et al. Ivacaftor in Infants Aged 4 to <12 Months with Cystic Fibrosis and a Gating Mutation. Results of a Two-Part Phase 3 Clinical Trial. Am J Respir Crit Care Med. 2021;203(5):585–93.

18. Ramsey BW, Davies J, McElvaney NG, Tullis E, Bell SC, Dřevínek P. A CFTR potentiator in patients with cystic fibrosis and the G551D mutation. N Engl J Med. 2011;365(18):1663–72.

19. Rosenfeld M, Wainwright CE, Higgins M, Wang LT, McKee C, Campbell D, et al. Ivacaftor treatment of cystic fibrosis in children aged 12 to <24 months and with a CFTR gating mutation (ARRIVAL): a phase 3 single-arm study. Lancet Respir Med. 2018;6(7):545–53.

20. Keating D, Marigowda G, Burr L, Daines C, Mall MA, Mckone EF, et al. VX-445–Tezacaftor– Ivacaftor in Patients with Cystic Fibrosis and One or Two Phe508del Alleles. New England Journal of Medicine. 2018;379(17):1612–20.

21. Griese M, Costa S, Linnemann RW, Mall MA, Mckone EF, Polineni D, et al. Safety and Efficacy of Elexacaftor/Tezacaftor/Ivacaftor for 24 Weeks or Longer in People with Cystic Fibrosis and One or More F508del Alleles: Interim Results of an Open-Label Phase 3 Clinical Trial. American Journal of Respiratory and Critical Care Medicine. 2021;203(3):381–5.

22. Heijerman HGM, McKone EF, Downey DG, Van Braeckel E, Rowe SM, Tullis E, et al. Efficacy and safety of the elexacaftor plus tezacaftor plus ivacaftor combination regimen in people with cystic fibrosis homozygous for the F508del mutation: a double-blind, randomised, phase 3 trial. Lancet. 2019.

23. Middleton PG, Mall MA, Drevinek P, Lands LC, McKone EF, Polineni D, et al. Elexacaftor-Tezacaftor-Ivacaftor for Cystic Fibrosis with a Single Phe508del Allele. N Engl J Med. 2019;381(19):1809–19.

24. Nichols DP, Paynter AC, Heltshe SL, Donaldson SH, Frederick CA, Freedman SD, et al. Clinical Effectiveness of Elexacaftor/Tezacftor/Ivacaftor in People with Cystic Fibrosis. Am J Respir Crit Care Med. 2021.

25. Zemanick ET, Taylor-Cousar JL, Davies J, Gibson RL, Mall MA, McKone EF, et al. A Phase 3 Open-Label Study of Elexacaftor/Tezacaftor/Ivacaftor in Children 6 through 11 Years of Age with Cystic Fibrosis and at Least One F508del Allele. Am J Respir Crit Care Med. 2021;203(12):1522–32.

26. Hutchinson I, McNally P. Appearance of Pancreatic Sufficiency and Discontinuation of Pancreatic Enzyme Replacement Therapy in Children with Cystic Fibrosis on Ivacaftor. Ann Am Thorac Soc. 2021;18(1):182–3.

27. Nichols AL, Davies JC, Jones D, Carr SB. Restoration of exocrine pancreatic function in older children with cystic fibrosis on ivacaftor. Paediatr Respir Rev. 2020.

28. Gelfond D, Heltshe S, Ma C, Rowe SM, Frederick C, Uluer A, et al. Impact of CFTR Modulation on Intestinal pH, Motility, and Clinical Outcomes in Patients With Cystic Fibrosis and the G551D Mutation. Clin Transl Gastroenterol. 2017;8(3):e81.

29. Ooi CY, Syed SA, Rossi L, Garg M, Needham B, Avolio J, et al. Impact of CFTR modulation with Ivacaftor on Gut Microbiota and Intestinal Inflammation. Sci Rep. 2018;8(1):17834.

30. Goralski JL, Hoppe JE, Mall MA, McColley SA, McKone E, Ramsey B, et al. Phase 3 Open-Label Clinical Trial of Elexacaftor/Tezacaftor/Ivacaftor in Children Aged 2 Through 5 Years with Cystic Fibrosis and at Least One F508del Allele. Am J Respir Crit Care Med. 2023.

31. Schwarzenberg SJ, Vu PT, Skalland M, Hoffman LR, Pope C, Gelfond D, et al. Elexacaftor/tezacaftor/ivacaftor and gastrointestinal outcomes in cystic fibrosis: Report of promise-GI. J Cyst Fibros. 2022.

32. Accurso FJ, Rowe SM, Clancy JP, Boyle MP, Dunitz JM, Durie PR, et al. Effect of VX-770 in persons with cystic fibrosis and the G551D-CFTR mutation. N Engl J Med. 2010;363(21):1991–2003.

33. Boon M, Claes I, Havermans T, Fornes-Ferrer V, Calvo-Lerma J, Asseiceira I, et al. Assessing gastro-intestinal related quality of life in cystic fibrosis: Validation of PedsQL GI in children and their parents. PLoS One. 2019;14(12):e0225004.

34. Hayee B, Watson KL, Campbell S, Simpson A, Farrell E, Hutchings P, et al. A high prevalence of chronic gastrointestinal symptoms in adults with cystic fibrosis is detected using tools already validated in other GI disorders. United European Gastroenterol J. 2019;7(7):881–8.

35. Quittner AL, Sweeny S, Watrous M, Munzenberger P, Bearss K, Gibson Nitza A, et al. Translation and linguistic validation of a disease-specific quality of life measure for cystic fibrosis. J Pediatr Psychol. 2000;25(6):403–14.

36. McCoy KS, Blind J, Johnson T, Olson P, Raterman L, Bai S, et al. Clinical change 2 years from start of elexacaftor-tezacaftor-ivacaftor in severe cystic fibrosis. Pediatr Pulmonol. 2023;58(4):1178–84.

37. Dabbous HK, Mohamed YAE, El-Folly RF, El-Talkawy MD, Seddik HE, Johar D, et al. Evaluation of Fecal M2PK as a Diagnostic Marker in Colorectal Cancer. J Gastrointest Cancer. 2019;50(3):442–50.

38. Nichols AL, Davies JC, Jones D, Carr SB. Restoration of exocrine pancreatic function in older children with cystic fibrosis on ivacaftor. Paediatr Respir Rev. 2020;35:99–102.

39. Heijerman HGM, McKone EF, Downey DG, Van Braeckel E, Rowe SM, Tullis E, et al. Efficacy and safety of the elexacaftor plus tezacaftor plus ivacaftor combination regimen in people with cystic fibrosis homozygous for the F508del mutation: a double-blind, randomised, phase 3 trial. Lancet. 2019;394(10212):1940–8.

40. Ooi CY, Syed SA, Rossi L, Garg M, Needham B, Avolio J, et al. Impact of CFTR modulation with Ivacaftor on Gut Microbiota and Intestinal Inflammation. Scientific reports. 2018;8.

41. Hisert KB, Heltshe SL, Pope C, Jorth P, Wu X, Edwards RM, et al. Restoring Cystic Fibrosis Transmembrane Conductance Regulator Function Reduces Airway Bacteria and Inflammation in People with Cystic Fibrosis and Chronic Lung Infections. Am J Respir Crit Care Med. 2017;195(12):1617–28.

42. Mainz JG, Arnold C, Wittstock K, Hipler UC, Lehmann T, Zagoya C, et al. Ivacaftor Reduces Inflammatory Mediators in Upper Airway Lining Fluid From Cystic Fibrosis Patients With a G551D Mutation: Serial Non-Invasive Home-Based Collection of Upper Airway Lining Fluid. Front Immunol. 2021;12:642180.

43. Rosenfeld M, Wainwright CE, Higgins M, Wang LT, McKee C, Campbell D, et al. Ivacaftor treatment of cystic fibrosis in children aged 12 to <24 months and with a CFTR gating mutation (ARRIVAL): a phase 3 single-arm study. Lancet Respir Med. 2018;6(7):545–53.

44. Rosenfeld M, Cunningham S, Harris WT, Lapey A, Regelmann WE, Sawicki GS, et al. An open-label extension study of ivacaftor in children with CF and a CFTR gating mutation initiating treatment at age 2-5 years (KLIMB). J Cyst Fibros. 2019;18(6):838–43.

45. Hebestreit H, Sauer-Heilborn A, Fischer R, Kading M, Mainz JG. Effects of ivacaftor on severely ill patients with cystic fibrosis carrying a G551D mutation. J Cyst Fibros. 2013;12(6):599–603.

46. Ooi C, Garg M, Young K, Needham B, Leach S, Jaffe A, et al. Improvement in Intestinal Inflammation on Ivacaftor. 2016;51:272-.

47. Chung-Faye G, Hayee B, Maestranzi S, Donaldson N, Forgacs I, Sherwood R. Fecal M2- pyruvate kinase (M2-PK): a novel marker of intestinal inflammation. Inflamm Bowel Dis. 2007;13(11):1374–8.

48. Pang T, Leach ST, Katz T, Jaffe A, Day AS, Ooi CY. Elevated fecal M2-pyruvate kinase in children with cystic fibrosis: a clue to the increased risk of intestinal malignancy in adulthood? Journal of gastroenterology and hepatology. 2015;30(5):866–71.

49. Rathod B, Chak S, Patel S, Shard A. Tumor pyruvate kinase M2 modulators: a comprehensive account of activators and inhibitors as anticancer agents. RSC Med Chem. 2021;12(7):1121–41.

50. Maisonneuve P, Marshall BC, Knapp EA, Lowenfels AB. Cancer risk in cystic fibrosis: a 20-year nationwide study from the United States. J Natl Cancer Inst. 2013;105(2):122–9.

51. Kapoor S, Chatterjee DR, Chowdhury MG, Das R, Shard A. Roadmap to Pyruvate Kinase M2 Modulation - A Computational Chronicle. Curr Drug Targets. 2023.

52. Birch RJ, Peckham D, Wood HM, Quirke P, Konstant-Hambling R, Brownlee K, et al. The risk of colorectal cancer in individuals with mutations of the cystic fibrosis transmembrane conductance regulator (CFTR) gene: An English population-based study. J Cyst Fibros. 2022.

53. Amaral MD, Quaresma MC, Pankonien I. What Role Does CFTR Play in Development, Differentiation, Regeneration and Cancer? Int J Mol Sci. 2020;21(9).

