## Supplementary information for "Reduction in abdominal symptoms (CFAbd-Score), faecal M2-pyruvate-kinase and Calprotectin over one year of treatment with Elexacaftor-Tezacaftor-Ivacaftor in people with CF aged ≥12 years – The RECOVER study"

### The CFAbd-Score

The latest CFAbd-Score version as validated according to FDA guidelines for development of a PROM as published 2019 (1) consists of 28 items grouped into five generic domains: “pain symptoms“ (four items), “disorders of bowel movement“ (eight items), “disorders of appetite“ (five items), “gastroesophageal reflux disease“ (GERD) symptoms (three items), and “quality of life impairment“ (QoL, eight items). CFAbd-Score items refer to a preceding 2-week recall period. Responses for 22 items are registered on a six-point Likert scale specifying either symptom severity ranging from “no problem” to “problem is as bad as it can be”, or symptom frequency ranging from “not at all/never” to “always/daily”. Pain intensity is rated on a scale from 0 (“no pain”) to 10 (“worst pain imaginable”) with mood-expressing faces, and pain duration is rated on a time scale.

Furthermore, among its 28 items, the CFAbd-Score includes questions regarding bowel movement and stool characteristics, i.e. “flatulence”, “constipation”, “fatty stools”, “foul-smelling stools”, “stool consistency”, “stool color”, (modified Bristol Stool Chart), “daily stool frequency” and “pain during bowel movements”, as qualitative work and clinical experience indicated their importance. (1-3)

Other items included in the latest version of the CFAbd-Score are: “abdominal pain”, “abdominal pain intensity”, “abdominal pain duration”, “lack of appetite”, “forced feeding”, “taste loss”, “bloating”, “nausea”, “heartburn”, “reflux” and “vomiting”, as well as items related to psychosocial factors such as: “reduced concentration”, “reduced physical activity”, “reduced productivity”, “fatigue”, “frustration”, “sadness”, “night waking”, “sleep difficulties” and “embarrassment”.

Given the intrinsic complexity of CF symptomatology, scientific evaluation of CFAbd-Scores requires the use of a specific algorithm developed during the distinct development and validation stages of the CFAbd-Score. This algorithm calculates a total score as well as sub-scores associated with the above-mentioned domains by assigning different weights to different items and domains to ensure optimal sensitivity. Accordingly, completed questionnaires resulting from international collaborations are always centrally scored at the CF Center in Brandenburg an der Havel, Germany.

### References


1. Jaudszus A, Zeman E, Jans T, Pfeifer E, Tabori H, Arnold C, et al. Validity and Reliability of a Novel Multimodal Questionnaire for the Assessment of Abdominal Symptoms in People with Cystic Fibrosis (CFAbd-Score). Patient. 2019;12(4):419-28.
2. Tabori H, Arnold C, Jaudszus A, Mentzel HJ, Renz DM, Reinsch S, et al. Abdominal symptoms in cystic fibrosis and their relation to genotype, history, clinical and laboratory findings. PLoS One. 2017;12(5):e0174463.
3. Tabori H, Jaudszus A, Arnold C, Mentzel HJ, Lorenz M, Michl RK, et al. Relation of Ultrasound Findings and Abdominal Symptoms obtained with the CFAbd-Score in Cystic Fibrosis Patients. Sci Rep. 2017;7(1):17465.
